# Microscopic Review of Nasopharyngeal Swabs as a Means of Benchside Quality Assurance

**DOI:** 10.1101/2020.04.06.20050088

**Authors:** AT. Matelski, N. Siddique, P. Mead, T. Raimondi, L. Jorgenson, O. Calderon, K. Harman, JJ. Farrell

**Affiliations:** University of Illinois College of Medicine, Peoria, IL; OSF System Laboratory, OSF St. Francis Medical Center, Peoria, IL

## Abstract

Nasopharygneal swabs (NPS) are the collection modality recommended by the Centers for Disease Control and Prevention (CDC) for reverse-transcription polymerase chain reaction (RT-PCR) testing for SARS-CoV2. NPS gather both extracellular material and human respiratory epithelial cells and, when used with RT-PCR, have reliable sensitivity for detection of viral infection. However, at our institution, we identified a 1.7% re-order rate within 7-days for NPS for respiratory pathogen RT-PCR, which we hypothesize may be due to low confidence in adequate sample collection. We sought to identify an inexpensive and accessible strategy for benchside quality assurance of NPS adequacy by observing microscopic content of viral transport media. For 801 NPS samples collected in November 2019, we air-dried and safranin-stained aliquots of viral transport media on glass slides. We then counted morphologically distinct ciliated columnar epithelial cells (CCEs). 19% of samples negative by RT-PCR for respiratory pathogens had no CCEs, while just 6% of positive samples exhibited the same. Pearson’s Chi-squared test was used to compare presence of CCEs between samples that were positive and negative for respiratory pathogens by RT-PCR *(p=3*.*3×10*^*-38*^). The negative predictive value (NPV) of finding no identifiable CCEs on microscopy was 85%. We posit that samples without identifiable CCEs may have been inadequately collected. The basic, benchside protocol using inexpensive laboratory reagents that we describe here could help improve accuracy and accessibility of NPS and RT-PCR testing for SARS-CoV2 and other respiratory pathogens while conserving limited resources in the face of a pandemic.

## Introduction

The emergence of the novel coronavirus SARS-CoV2 in late 2019 that has since been declared a pandemic by the WHO has placed increased pressure on identifying testing modalities that are readily available and reliable. Initial identification of the causative pathogen by the China CDC was accomplished via recovery of bronchoalveolar lavage fluid and subsequent RT-PCR testing. The investigative team then inoculated human respiratory epithelial cells in culture and identified the same pathogen via next-generation sequencing of the supernatant and direct observation of viral cytology via electron microscopy.^1^

The tropism of SARS-CoV2 for human respiratory epithelial cells, also called ciliated columnar epithelial cells (CCEs), is similar to many respiratory viruses.^2^ Nasopharyngeal swabs (NPS), which gather both extracellular and human cellular material from the upper airway, capture viral genetic material. NPS are the preferred collection modality for respiratory samples undergoing pathogen detection via multiplex PCR when testing for respiratory viruses.

The CDC currently recommends NPS for collection of specimens for persons under investigation for COVID-19 in the U.S. and suggests that collection of oropharyngeal swabs or sputum is of lower priority.^3^ Previous studies have suggested that viral loads of SARS-CoV2 may be higher in the nose than in the throat.^4^ NPS have demonstrated greater sensitivity than oropharyngeal swabs for both common viruses and SARS-CoV2.^5-8^ NPS use is straightforward and preferred by frontline healthcare personnel, and it does not carry the potential to generate bioaerosol as a bronchoalveolar lavage might. ^9,10^ These features are especially relevant during a pandemic, when widespread and accessible testing is an essential pillar of the public health response.

However, given the technique-dependent nature of collection and patient discomfort associated with NPS, it is recognized that inadequate sample collection alters accuracy.^11-13^ This phenomenon is also recognized among patients with COVID-19. A patient in a Thai hospital in January 2020 had NPS and oropharyngeal swabs negative for SARS-CoV2 but subsequent RT-PCR testing of bronchoalveolar lavage fluid was positive for the virus.^14^ Three patients in Zhangjiang, China had intermittently negative throat swabs despite confirmed COVID-19.^15^ Nasopharyngeal swabbing was recommended as a co-test among other specimen sites due to a high false negative rate (37%) among 398 NPS collected in Beijing.^8^ Acquiring adequate nasopharyngeal swabs is a clear priority for accurate diagnosis of COVID-19.

In our institutional laboratory, 11,940 total NPS samples were submitted between Sep 2017 and June 2019 for respiratory pathogen RT-PCR. We analyzed patients who received a second swab within 7 days of the original; in total, there were 198 repeat swabs (1.7%), which we hypothesized may have been ordered due to lack of clinician confidence in initial NPS.

Previous studies have shown that cellular content of NPS is key for sensitivity in respiratory pathogen identification by RT-PCR, but methodology was often expensive and complex.^13,16,17^ Prior to the beginning of the SARS-CoV2 outbreak, we conducted an investigation of a simple method for identifying cellular content as a surrogate for sample adequacy. Now, these data have potentially important implications for the public health response to SARS-CoV2, as a means of improving NPS accuracy and conserving limited testing resources.

## Materials & Methods

We processed all NPS samples destined for multiplex respiratory pathogen RT-PCR or influenza RT-PCR testing in our system laboratory during the month of November 2019 (n=801). Review of these samples was approved by the University of Illinois College of Medicine-Peoria institutional review board (IRB) prior to initiation. All samples were collected as part of routine patient care in viral transport media (*BD, Franklin Lakes, NJ, USA)*. They originated from 286 unique patients: 50% were female and median age was 34. Samples were processed, on average, 4.3 days post-collection. Samples were vortexed for 3 seconds, 25 µL of media were pipetted onto a glass slide, and slides were air dried. Each was safranin stained for 1 minute, rinsed, and air dried. All slides were prepared by the same individual.

Under light microscope, each slide was reviewed at 40x. Each sample was examined for ciliated columnar epithelial cells (CCEs), which have a distinct morphology and can be clearly identified at 40x (see Figure 1). A group of medical students, residents, and laboratory technicians were trained to recognize CCEs during a single didactic session. These individuals independently counted CCEs across 3 randomly selected fields and averaged the result. Each sample was reviewed twice by independent reviewers.

**Figure 1.**
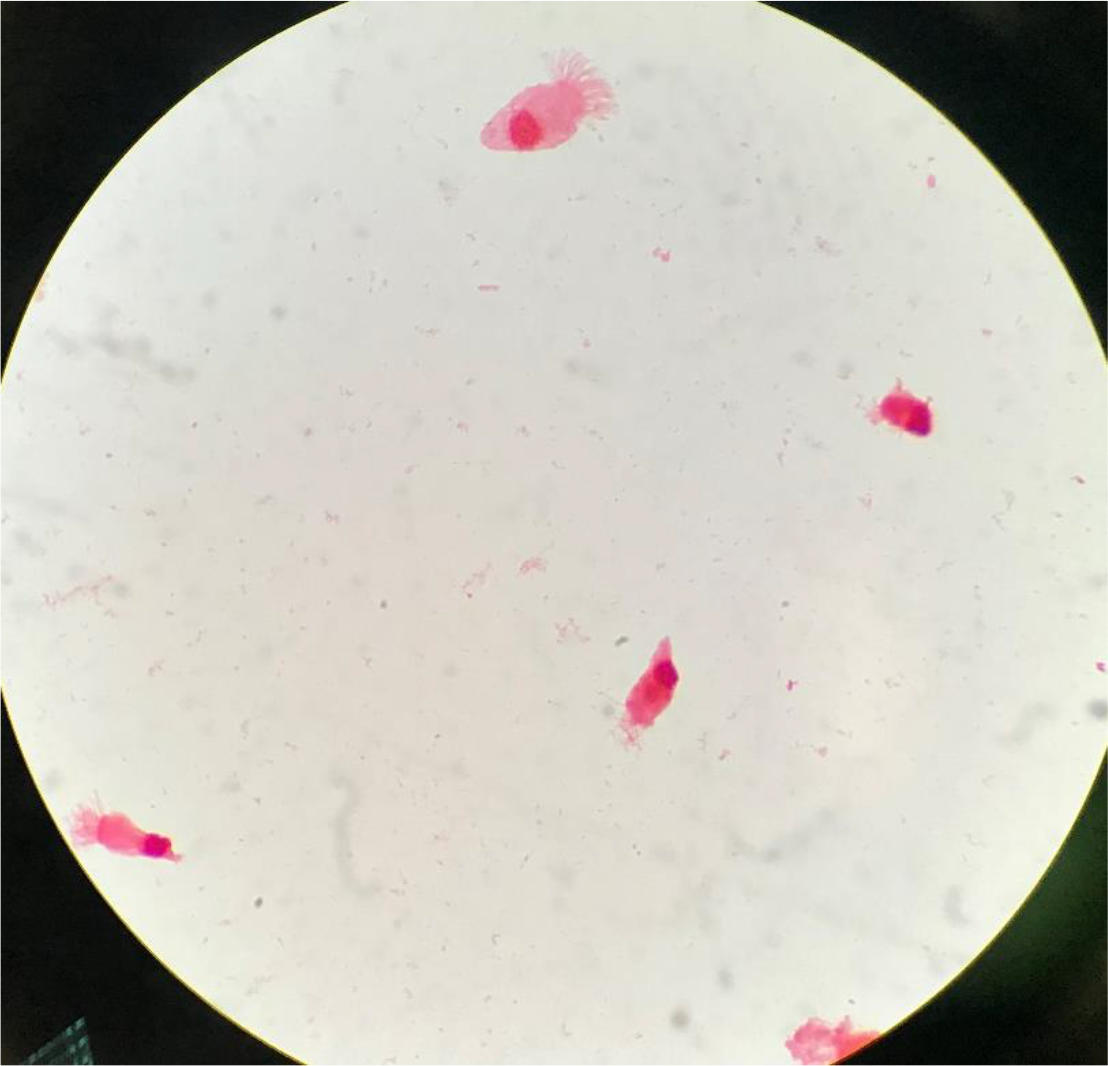
Safranin-stained ciliated columnar epithelial cells (CCEs) as seen at 40x microscopy.

Inter-rater reliability was determined using a Bland-Altman plot of reviewer 3-field averages. A correlation coefficient was calculated for average number of CCEs/hpf and age. Average number of CCEs/hpf were compared between samples that were positive and negative for respiratory pathogens by RT-PCR using Pearson’s Chi-squared test. Negative predictive value (NPV) was calculated for samples without identifiable CCEs.

## Results

There were 517 negative samples and 284 positive samples as verified by RT-PCR for respiratory pathogens. There was no statistically significant difference between positive and negative samples for sex, but they did vary meaningfully for age, with an average of 22 years for negative samples and 44 years for positive samples (*p<0*.*01*) (Table 1). Despite this statistically significant difference in patient age between positive and negative samples, identifiable CCEs were not correlated with patient age (R^2^_positive samples_ = 0.01; R^2^ _negative samples_ =0.01) (Figure 2).

**Table 1.**

|  | Positive |  | Negative |  |
| --- | --- | --- | --- | --- |
|  | Number | % | Number | % |
| Female | 134 | 47% | 264 | 51% |
| Age, Years (Average) | 21.9 |  | 43.9 |  |
| Age, Years (Median) | 5 |  | 53 |  |

**Figure 2.**
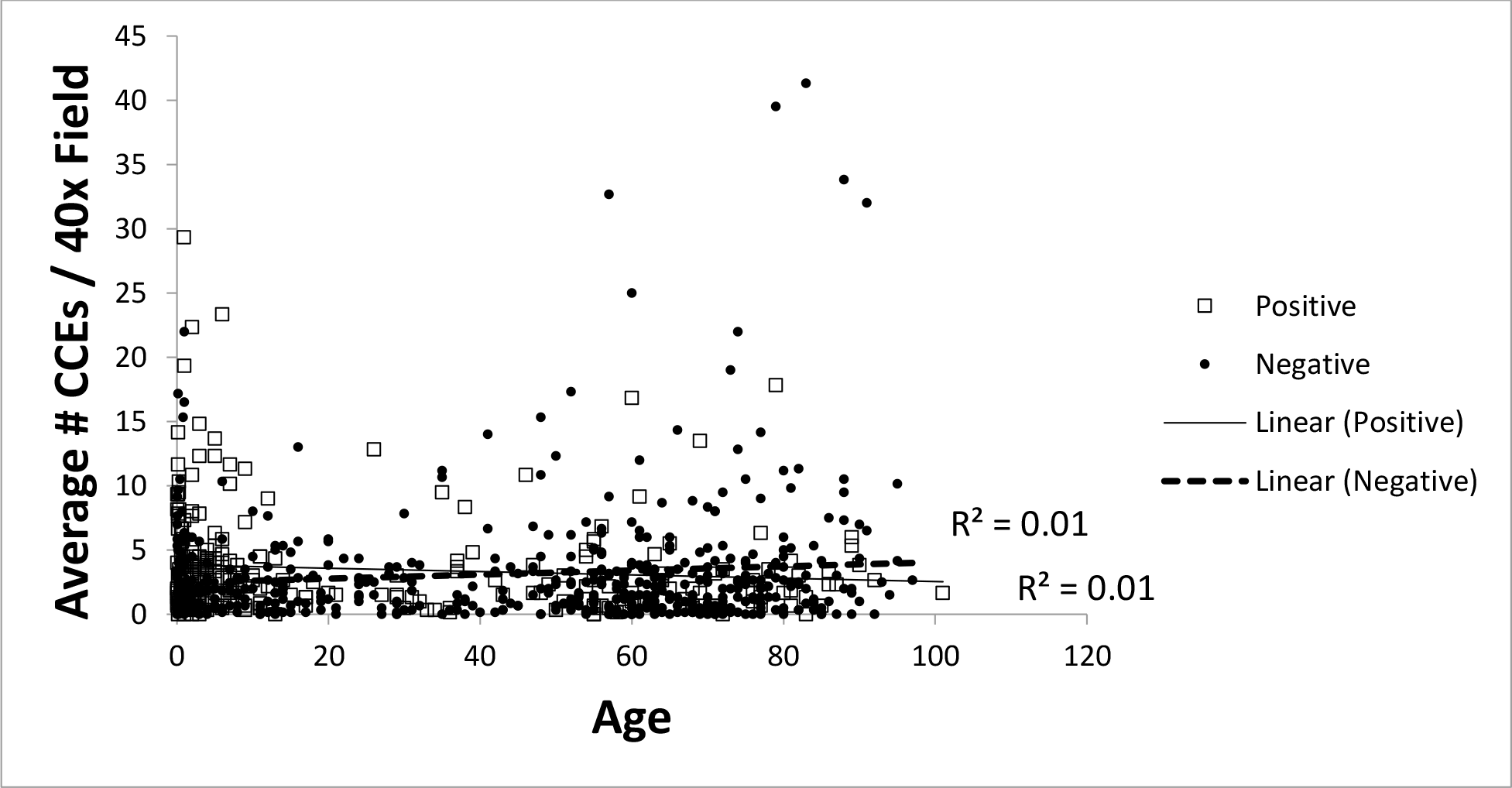

The Bland-Altman plot for inter-rater reliability determined that 96.5% of inter-rater difference was within two standard deviations (average difference = 2.0 CCEs; SD = 6.1). The majority of both positive and negative samples had at least one CCE/hpf, with an average of 3.7 CCEs/hpf in positive samples and 3.2 CCEs/hpf in negative samples (*p=0*.*18*). However, 19% of negative samples had no CCEs visible on microscopy, while just 6% of positive samples exhibited the same (Table 2). The Chi-Squared test comparing CCEs/hpf between positive and negative samples was 3.3 × 10^−38^ (*α=0*.*05*). There were 100 negative samples without identifiable CCEs out of 118 total samples without identifiable CCEs, which yields a NPV of 85%.

**Table 2.**
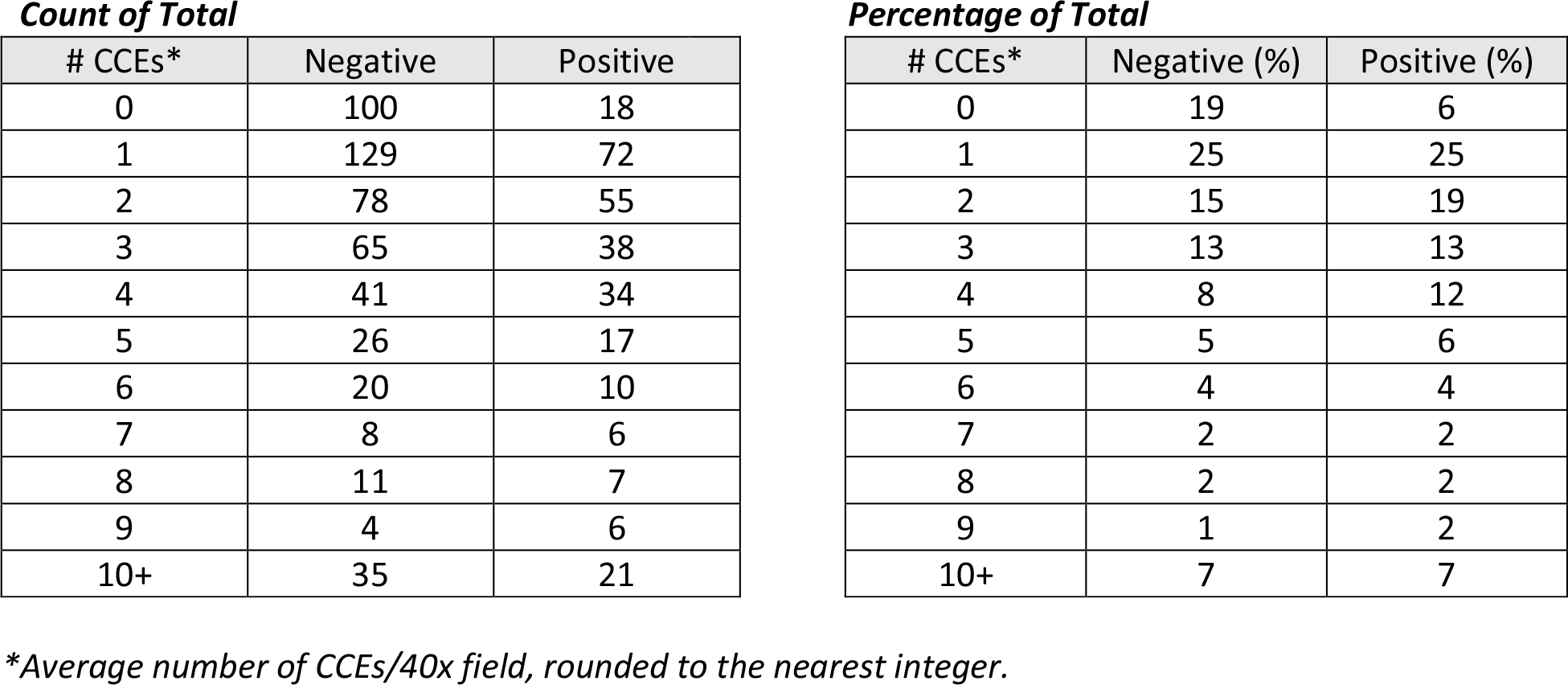

| *Count of Total* |  |  | *Percentage of Total* |  |  |
| --- | --- | --- | --- | --- | --- |
| # CCEs* | Negative | Positive | # CCEs* | Negative (%) | Positive (%) |
| 0 | 100 | 18 | 0 | 19 | 6 |
| 1 | 129 | 72 | 1 | 25 | 25 |
| 2 | 78 | 55 | 2 | 15 | 19 |
| 3 | 65 | 38 | 3 | 13 | 13 |
| 4 | 41 | 34 | 4 | 8 | 12 |
| 5 | 26 | 17 | 5 | 5 | 6 |
| 6 | 20 | 10 | 6 | 4 | 4 |
| 7 | 8 | 6 | 7 | 2 | 2 |
| 8 | 11 | 7 | 8 | 2 | 2 |
| 9 | 4 | 6 | 9 | 1 | 2 |
| 10+ | 35 | 21 | 10+ | 7 | 7 |

*\*Average number of CCEs/40x field, rounded to the nearest integer.*

## Discussion

This study demonstrated a meaningful difference in identifiable CCEs on microscopic review between NPS that were negative or positive by RT-PCR for respiratory pathogens. There was greater association between lack of identifiable CCEs and RT-PCR negative NPS, which we posit is partially attributable to inadequately collected swabs. This feature also demonstrated a useful discriminatory NPV. Given this finding, microscopic review of CCEs using simple, inexpensive reagents and equipment could be a helpful step in the diagnostic algorithm for upper respiratory viral infection. Situations in which there is high clinical suspicion but a negative RT-PCR may benefit from rapid microscopic review using a streamlined, benchside version of this protocol, which could save limited resources like NPS, viral transport media, and RT-PCR kits.

Given that CCEs could not be identified in 6% of positive samples, it is plausible that there was imperfect slide preparation, which resulted in loss of cellular content, or that pipetting from the viral media did not reliably collect CCEs that may have been present on the rayon swab or in sediment. It is also plausible that in those cases of positive samples without CCEs, viral genetic material might be identifiable by RT-PCR without co-presence of CCEs, but the statistically significant comparative number of negative samples without identifiable CCEs makes this unlikely as the sole explanation.

This study is limited by being conducted a single-site with the attendant possibility of idiosyncratic collection techniques. It also did not have the granularity to assess variation of CCEs by pathogen type. Additionally, a set indication for NPS was not proscribed and providers may have used variable clinical criteria when ordering a test.

Having additional laboratory methods for quality assurance may be helpful in ensuring readily available and reliable testing. Using inexpensive laboratory reagents and light microscopy to identify CCEs has reasonable discriminatory capability and can help ensure adequate NPS for the diagnosis of COVID-19 and other respiratory viral illness. The basic, benchside protocol described here found the NPV of an absence of CCE on light microscopy was 85%, which could help improve accuracy and accessibility of NPS and RT-PCR testing for SARS-CoV2 and other respiratory pathogens while conserving limited resources in the face of a pandemic.

## Data Availability

Data can be made available upon publication in a peer-reviewed journal.

## Acknowledgements

This research received no specific grant from any funding agency in the public, commercial, or not-for-profit sectors. However, we would like to thank the staff of the OSF system microbiology and immunology labs for their unwavering support, especially Heather Shaner, Keirsten Grindler and Jacquelyn Lynn.

